# Mixed-effects polygenic risk score Phenome-wide association study detects genetic correlation between colorectal cancer risk and phenotype data extracted from the electronic health record

**DOI:** 10.1101/2025.02.26.25322864

**Authors:** Elisabeth A. Rosenthal, Wei-Qi Wei, Yuan Luo, Bahram Namjou-Khales, Daniel J. Schaid, Edward D. Esplin, Michael Lape, Leah Kottyan, Jennifer Allen Pacheco, Chunhua Weng, Adam Samuel Gordon, Iftikhar J. Kullo, David R. Crosslin, William M. Grady, Li Hsu, Ulrike Peters, Gail P. Jarvik

## Abstract

Many factors, including environmental and genetic variables, contribute to Colorectal Cancer (CRC) risk. Some of these risk factors may share underlying genetics with CRC. We investigated potential shared genetics by performing a Phenome-wide association study (PheWAS) with a multi-ancestry CRC polygenic risk score (PRS). The discovery cohort (N=426,464) consisted of ancestrally diverse participants from the United Kingdom Biobank. The replication cohort (N=87,271) consisted of ancestrally diverse participants from the electronic Medical Records and Genomics Network. We used a mixed-effects model to adjust for the presence of related individuals in both datasets. To preserve power, we limited testing to ancestor phecodes derived from the electronic health record (EHR), which were not likely to be a result of CRC or its treatment. We discovered and replicated associations between the CRC PRS and breast cancer, prostate cancer, obesity, smoking and alcohol use (discovery p< 1.1e-4; replication p<0.0019). As these results corroborate findings from other studies using orthogonal methods, we demonstrate that a CRC PRS can be used as a proxy for genetic risk for CRC when investigating shared genetics between CRC and other phenotypes. Further study of the relationship between PRS from multiple traits with EHR data may reveal additional shared genetic factors.

## Introduction

Colorectal cancer (CRC) is the second most deadly cancer in the United States, with an average mortality of 13%, and increasing incidence among younger individuals (Siegel et al. 2023). Risk of CRC is complex and known to be associated with both environmental and genetic risk factors. Associated environmental factors for CRC risk include smoking, diet, alcohol intake, and physical activity, as well as the comorbidities obesity and diabetes (Sawicki et al. 2021; Jiang et al. 2011; Pearson-Stuttard et al. 2021; Kyrgiou et al. 2017). Genetic heritability of CRC is estimated to be between 12 and 40% (Jiao et al. 2014; Graff et al. 2017). Several genes are known to underlie high monogenic risk, but these account for only ~20% of familial cancers (Lowery et al. 2016; Patel and Ahnen 2012; Rosenthal et al. 2018). Polygenic risk scores (PRSs), which aggregate the effects of multiple small effects across the genome, may capture some of this missing heritability (Hatchell et al. 2019; Manolio et al. 2009). The heritability explained by common SNPs for CRC has been estimated to be between 7 and 11% accounting for ~73% of familial risk. (Fernandez-Rozadilla et al. 2023; Zhang et al. 2020; Jiao et al. 2014). In addition, PRS have been shown to predict CRC risk, independently of family history, indicating that they may also explain some portion of non-familial CRC (Mars et al. 2022; Tada et al. 2016; Archambault et al. 2020)). Moreover, CRC PRS have been shown to be more predictive for younger adults than older adults, indicating that they may be helpful in identifying individuals who would benefit from increased or earlier screening (Archambault et al. 2020).

Mendelian randomization (MR) and cross-trait linkage disequilibrium (LD) score regression provide evidence of shared genetics underlying risk for CRC and other traits. MR is a technique that can determine if risk factors are merely associated with an outcome of interest, or if they are causal for that outcome by using underlying genetics of the risk factor as instrumental variables (Burgess and Thompson 2013; Burgess, Dudbridge, and Thompson 2016). For example, although it is well known that body mass index (BMI) is positively associated with CRC risk, MR studies indicate that genetic predictors of BMI may be causally associated with CRC risk (Renehan et al. 2008; Kyrgiou et al. 2017; Bouras et al. 2024). Similarly, MR has been used to show a genetic correlation between early-onset CRC and alcohol consumption (Laskar et al. 2024). Traditional MR studies focus on a single gene, as increasing the number of SNPs involved can lead to an increased chance of violating instrumental variable assumptions, reducing the reliability of MR. Additionally, pleiotropy influencing multiple biological pathways (i.e., non-horizontal pleiotropy) violates the instrumental variable assumptions and can lead to misinterpretations of causal relationships (Burgess et al. 2017; Bowden et al. 2017). Alternatively, cross-trait LD score regression measures genetic correlation genome-wide between two traits (Lee et al. 2012; Bulik-Sullivan et al. 2015). For example, CRC has been shown to be genetically correlated with breast, lung, and esophageal cancers in individuals of European ancestry (Lindström et al. 2023). Similarly, CRC has been shown to be genetically correlated with fasting insulin, BMI, and smoking using LD-score regression (Fernandez-Rozadilla et al. 2023).

Here, we investigate the genetic correlation underlying CRC risk and other phenotypes by performing PheWAS with a CRC PRS (Carroll, Bastarache, and Denny 2014). The CRC PRS used here has been developed in individuals of European and Asian ancestry, and validated in multiple datasets (Thomas et al. 2023). We used participant data from the United Kingdom Biobank (UKBB) as a discovery cohort and participant data from the electronic Medical Records and Genomics (eMERGE) study as a replication cohort. As biobanks often include related individuals, we used a mixed model analysis, accounting for possible phenotype correlation among related individuals, allowing efficient use of all data. We corroborate the shared genetic relationship between CRC and multiple traits, such as breast cancer, obesity, smoking and alcohol use disorder. We also find evidence for potential shared genetics underlying CRC risk and prostate cancer.

## Methods

### Participants

The discovery cohort was derived from the UKBB (Bycroft et al. 2018), extracted on September 18, 2023. We included participants for whom electronic health record (EHR) data was available and for whom the PRS could be calculated. In addition to the EHR data, we extracted cancer registry and death registry data. We then excluded any participants who developed cancer before age 18, resulting in a sample size of N=426,464 (N female = 232,979, N male = 193,485) (Table 1).

**Table 1:** Percent female (%F), Age (mean and range), and total sample count by cohort. The replication cohort is also broken down by site and sorted by the number of adult participants at each site. UKBB=United Kingdom Biobank; eMERGE=electronic Medical Records and Genomics Network; CHOP=Children’s Hospital of Philadelphia; CCHMC= Cincinnati Children ‘s Hospital Medical Center; UW=University of Washington.

| COHORT | %F | Age | Total |
| --- | --- | --- | --- |
| **UKBB** | 54.6 | 65 (21,86) | 426,464 |
| **eMERGE** | 55.8 | 63 (18,90) | 87,271 |
| Boston Children's | 45.9 | 20 (18,25) | 133 |
| CHOP | 55.9 | 20 (18,36) | 699 |
| CCMC | 51.9 | 23 (18,64) | 1,346 |
| Columbia | 48.8 | 62 (19,89) | 1,970 |
| Geisinger | 47.4 | 69 (23,89) | 3,103 |
| Kaiser-UW | 56.9 | 83 (54,89) | 3,283 |
| Northwestern | 82.7 | 60 (19,90) | 4,708 |
| Marshfield Clinic | 60.5 | 78 (32,90) | 4,756 |
| Mt. Sinai | 59.3 | 58 (20,89) | 6,167 |
| Mayo Clinic | 48.2 | 70 (18,89) | 9,133 |
| Vanderbilt | 54.9 | 63 (18,90) | 21,411 |
| Harvard | 54.6 | 59 (18,89) | 30,562 |

The replication cohort was derived from adult participants in the eMERGE phase 3 study (N = 87,271; N female = 48,737, N male = 38,534), as described previously (Stanaway et al. 2019)(Table 1). The eMERGE phase 3 network consisted of 12 sites across the continental United States. Data for these participants were extracted from the EHR through January 2022. The 12 sites in the eMERGE cohort included the following clinical site types: hospital or primary care, adult or pediatric, and specialty clinics.

### Genotype data and PRS calculation

Methods for genotype data collection for both UKBB and eMERGE participants have been published previously (Stanaway et al. 2019; Bycroft et al. 2018). SNP data was extracted from the imputed genotypes in UKBB (Resource 530; Category 100319) and eMERGE. The genotype data in eMERGE was harmonized and combined over several genotype arrays and then imputed genome-wide. We used the principal components of ancestry (UKBB Data-Field 22009) and kinship estimates (UKBB Data-Field 22021) provided by the UKBB. We calculated principal components of ancestry and estimated kinship in eMERGE, adjusting for recent family history, using the R package GENESIS v2.20.1 (Gogarten et al. 2019; R Core Team 2024) (see <u>Supplemental Material</u>). The allele effects for each SNP in the PRS were obtained from (Thomas et al. 2023) and can also be found in the PGS catalog (PGS003852). The PRS was calculated using the R package bigsnpr v1.10.8 (Privé et al. 2018), which accounts for potential strand reversal and allele mismatches.

### PheWAS Discovery

We transformed the international Classification of Diseases (ICD) 9/10 data from the medical records to phecodes using the R package PheWAS v0.99.6-1 with phecode map v1.2024 (Carroll, Bastarache, and Denny 2014; Wei et al. 2017; Wu et al. 2019). We defined the medical record to be the compilation of data from the EHR, death records (UKBB participants only), and cancer registries. In the situation where the death record contained more than one ICD code, we kept only the ICD code for the primary cause of death. For each phecode, a participant was assigned case status if that phecode was observed at least twice and was assigned control status if that phecode was never observed. Participants with exactly one observation of the phecode were excluded from analysis for that phecode. Therefore, the sample size varies by phecode. The phecode system is hierarchical, similar to the ICD system, so that ancestor codes define a broad phenotype, and child codes define increasingly specific phenotypes related to the ancestor. Therefore, a participant who is a case for a child code would also be a case for all of its ancestor codes. Due to this hierarchical nature, the number of cases is always highest for the ancestor codes relative to the child codes.

We performed a mixed model PheWAS, which accounts for kinship among the participants. We used the mixed model analysis from the R package GENESIS v2.32 to evaluate the association between the PRS and case/control status for each phecode, adjusting for sex, age, and the first four principal components of ancestry (Gogarten et al. 2019). Case/control status were encoded as factors (1, 0 respectively) and stored in VCF format in order to use the functionality of the R package GENESIS. Age was defined as the oldest age recorded in the medical record or age at death, when available. We excluded males from the analysis of breast cancer as it is rare in males, and we excluded females in the analysis of prostate cancer.

As many phecodes are correlated (e.g., ancestor phecodes and their child phecodes), using a Bonferonni p-value correction adjusting for all phecodes would be too conservative. Therefore, we focused our analysis on the ancestor codes (N = 510). As the PRS was derived with UKBB data as part of the training data, we expect significance for CRC ancestor phecodes (153, 565) and do not count them toward the number of tests. In addition, we removed all phecodes from the discovery analysis which could be a consequence of CRC or its treatment (N = 53) (e.g., Chemotherapy, Ileostomy status) (see <u>Supplemental Table</u> 1). Therefore the p-value cutoff used in the initial association was 0.05/455=1.1e-4.

### Replication

We counted the number of significant tests from the discovery analysis as the total number of tests for the replication analysis. Therefore, the p-value cutoff was determined as 0.05/N, where N is the number of significant ancestor phecodes, from UKBB, that are not likely to be a consequence of CRC or its treatment. We performed a mixed model PheWAS adjusting for age, sex, site, and the first four principal components of ancestry. We included site because ICD code usage varied across the twelve sites, depending on the type of clinic (hospital vs. primary care; adult vs. pediatric; general vs. specialty). As a supplementary analysis, we performed a mixed model PheWAS in the UKBB cohort, excluding participants who were coded as cases for CRC (phecode 153) for all phecodes that replicated in the eMERGE cohort. The purpose of this analysis was to detect possible bias in effect size estimates due to the presence of CRC cases in the initial discovery (see supplemental Material).

## Results

### Demographics

Females represented 55% and 56% of the participants in UKBB and eMERGE, respectively (Table 1). The average age was slightly higher in UKBB (65 ± 10.5 years) than eMERGE (63 ± 18 years). On average, in UKBB and eMERGE, females were two and four years younger than males, respectively. Additionally, the age distribution varied across sites in eMERGE. Younger participants were enrolled at Boston Children’s and Children’s Hospital of Philadelphia (CHOP), but the total sample size of adults (age >= 18) at these sites made up only < 1% of the total analyzed here. In eMERGE, the largest portion of participants (35%) were from Harvard, followed by Vanderbilt (25%). The majority of participants identified as White in both UKBB (94%) and eMERGE (80%). In UKBB 2.2% self-identified as Asian, 0.6% self-identified as Black, 2.3% self-identified as Other or Mixed ancestry, and 0.5% did not indicate a race. Similar to age, the self-identified race varied across eMERGE sites, with a low of 11% self-identifying as White at Mt. Sinai and up to 99.6% self-identifying as White at Geisinger. Overall, 8.1% self-identified as Black, with a range between 0.3% (Geisinger) and 65% (Mt. Sinai). Overall, 1% self-identified as Asian, with a range between 0% (Mt. Sinai) and 3.9% (Columbia). Only 0.1% self-identified as Native American or Pacific Islander, with the largest proportion (0.8%) at Kaiser/University of Washington (UW). Self-identified race was missing for 11% overall in eMERGE. Four percent of participants in eMERGE identified as Hispanic, with the majority at Harvard (1.8%) and Mt. Sinai (1.5%). There was no mechanism at eMERGE to self-identify as multiple races or ethnicities. However, there is genotypic variation and admixture among the participants in both cohorts (Figure 1).

**Figure 1:**
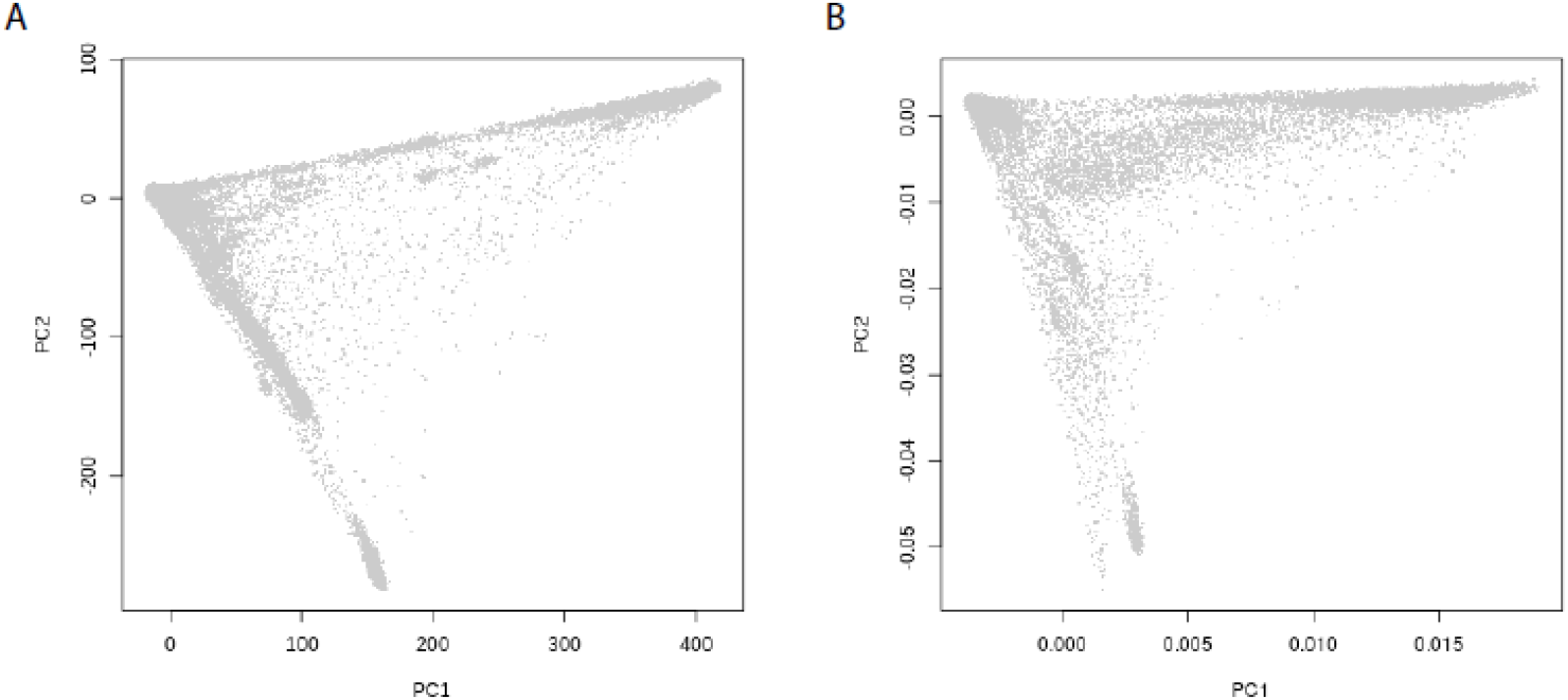
The first two principal components (PC) of ancestry in the A) Discovery and B) Replication cohorts. In both plots, the three points of the triangle, clockwise from the left-most point, represent European genetic ancestry, African genetic ancestry, and East Asian genetic ancestry.

### PRS

The overall mean and standard deviation (s.d.) of the PRS is 0.34 ±0.47 in UKBB and 0.61 ±0.47 in eMERGE. After adjusting for the first four PCs of ancestry, the mean was zero for both cohorts. The distribution of the ancestry adjusted PRS is not significantly different from a standard Normal distribution in both cohorts using a Kolmogorov-Smirnov test (p > 0.17), as expected.

### Discovery and Replication

The PRS was highly significantly associated with CRC in UKBB (p<2.2e-308), as expected (<u>Supplemental Table</u> 1). In addition, the PRS was associated with 22 ancestor phecodes that are likely a consequence of CRC or its treatment, such as Secondary malignant neoplasm and Chemotherapy (<u>Supplemental Table</u> 1). Excluding these phecodes, the PRS was associated with 26 ancestor phecodes at an alpha level of 1.1e-4 in UKBB (Table 2).

**Table 2:** Effect estimates (β) and p-values (p) for phecodes that are significantly associated with the CRC PRS in the discovery cohort (p< 1.1e-4) and the replication cohort (p<0.0019).

| Phecode | Description | Discovery |  | Replication |  |
| --- | --- | --- | --- | --- | --- |
| | | $\beta$ | p | $\beta$ | p |
| 208 | Benign neoplasm of colon | 0.246 | 2.2e-308 | 0.117 | 9.6e-169 |
| 175 | Acquired absence of breast | 0.05 | 3.3e-39 | 0.027 | 6.4e-04 |
| 1100 | Family history | 0.035 | 3.4e-27 | 0.03 | 4.3e-06 |
| 317 | Alcohol-related disorders | 0.044 | 3.3e-11 | 0.023 | 8.7e-04 |
| 174 | Breast cancer | 0.027 | 1.2e-09 | 0.028 | 1.5e-04 |
| 211 | Benign neoplasm of other parts of digestive system | 0.048 | 1.9e-09 | 0.051 | 1.0e-06 |
| 185 | Cancer of prostate | 0.022 | 2.2e-07 | 0.025 | 1.6e-03 |
| 038 | Septicemia | 0.026 | 2.5e-06 | 0.022 | 1.7e-04 |
| 278 | Overweight, obesity and other hyperalimentation | 0.016 | 5.4e-05 | 0.012 | 4.2e-04 |
| 318 | Tobacco use disorder | 0.024 | 9.8e-05 | 0.015 | 1.5e-04 |

The PRS was significantly associated with CRC in eMERGE (p=7.8e-65), as expected. Given that there were 26 significant ancestor phecodes that are not necessarily a consequence of CRC or its treatment in the UKBB data, we set the p-value cutoff to 0.05/26= 0.0019 for the replication. Ten of the ancestor phecodes replicated in eMERGE (Table 2): Septicemia; Family history; Breast Cancer; Acquired absence of breast; Cancer of prostate; Benign neoplasm of colon; Benign neoplasm of other parts of digestive system; Overweight, obesity and other hyperalimentation; Alcohol-related disorders; and Tobacco use disorder. Additionally, the direction of effect is the same across cohorts and the estimated effect sizes are similar across the cohorts. However, when we removed the CRC cases from the discovery UKBB cohort, the effect estimates for Septicemia and Acquired absence of breast reduced by approximately half (See Supplemental Material).

## Discussion

We provide evidence for genetic correlation between CRC and several traits, using the CRC PRS as a genetic score for CRC risk. The estimated effect size in the discovery and replication cohorts for these phecodes was similar across cohorts, indicating that the overall genetic correlation structure may be similar across the cohorts. Among non-CRC related cancers, only breast cancer in females and prostate cancer in males were found to be associated with the CRC PRS. Additionally, benign neoplasms of the colon and of other parts of the digestive system, which can become cancerous if not removed, were associated with the PRS. Unsurprisingly, we detected correlation between the PRS and Family History. Family History is defined as family history for any disorder, including neoplasms and cancer. It is likely that patients with CRC, prostate cancer, or breast cancer are asked for their family history in the clinic, which would explain this association.

We also detected an association between the CRC PRS and modifiable environmental phecodes believed to be risk factors for CRC. These include obesity, smoking, and alcohol use. Although we replicated the association with obesity, we did not replicate the association with diabetes as the p-value for diabetes (0.003) in the eMERGE replication cohort was slightly higher than the cutoff for replication (1.9e-3). We did not observe bias in the estimated effect sizes for these phecodes due to the presence of the CRC cases, indicating that the relationship between these behavioral risk factors may share underlying genetics with CRC risk. Conversely, although we detected an association with septicemia, there is evidence that the presence of CRC cases in the initial UKBB cohort may be biasing the estimated effect as the exclusion of these cases resulted in a change in effect size by half.

Our results replicate genetic correlations with CRC that were found in other studies using orthogonal methods. These include correlations with breast cancer, obesity, alcohol use, and smoking (Renehan et al. 2008; Kyrgiou et al. 2017; Bouras et al. 2024; Lindström et al. 2023; Fernandez-Rozadilla et al. 2023; Laskar et al. 2024). This indicates that using PRS to test for genetic correlations may be a complementary method to explore the joint genetic architecture of multiple traits, including traits that are thought to be behavioral, but have evidence of genetic causes (Polderman et al. 2015). We did not replicate the genetic correlation between CRC and lung (cancer within the respiratory system) or esophageal cancers that were previously reported (Lindström et al. 2023). It is possible that these were false positives, or that using a PRS is not as sensitive as cross-trait LD score regression, or there is lower power in the eMERGE dataset to detect an effect. We also detected association between CRC risk and prostate cancer, which has not been detected by the other methods, to our knowledge.

Interpretation of these results is limited by a few factors. First, the phecodes are built from ICD9/10 billing codes. Billing codes do not necessarily indicate the presence of a phenotype but rather that a clinical encounter related to that phenotype occurred (Wei et al. 2016). To reduce noise due to this issue, we considered participants as cases if they had the same phecode twice on separate clinical visits and as controls if there was no occurrence of that phecode. In addition, we used cancer registry data, which further improved the case definition for cancers. Moreover, we focused on the ancestor codes, which encompass a broader phenotype definition and therefore may be more coherent. For example, both diabetes I and II billing codes were observed for the same individuals (data not shown). Rather than focus on the descendant diabetes codes, we looked at the association between the CRC PRS and diabetes overall. Furthermore, billing code use is not consistent across clinics, which can result in a loss of power or increase type I error (Sulieman et al. 2022; Pendergrass and Crawford 2019). We observed this phenomenon in our data for vision phenotypes, necessitating adjustment by site (data not shown). Finally, behavioral conditions are not reliably recorded in EHR as billing codes (Pendergrass and Crawford 2019). Therefore, we may be undercounting the number of participants who smoke or use alcohol. However, smoking and alcohol behavior are typically assessed in the clinic (Adler and Stead 2015). As we observe an association between the PRS and both smoking and alcohol use in both cohorts, we believe this provides evidence for further investigation of a potential genetic correlation between CRC and smoking and alcohol consumption behaviors.

Another limitation is that the PRS was developed including data from the UKBB. Therefore, the PRS may be confounded with some phenotypes and behaviors within the UKBB. However, participants from the UKBB made up only 13.5% of the training data (4.8% of the CRC cases), reducing potential confounding (Thomas et al. 2023; Fernandez-Rozadilla et al. 2023). Similarly, participants from eMERGE were used to validate the PRS (27% of the total validation cohort). However, as part of the validation dataset, no adjustments were made to the PRS model. Therefore, it is unlikely that unintentional confounding between the PRS and phenotypes of interest are present due to the eMERGE participants.

Further studies assessing PheWAS for other trait PRS may provide evidence in support or against the correlations observed here. For example, PRSs exist for obesity and several cancers (Lennon et al. 2024). A multiple PRS PheWAS may reveal shared genetics across traits. Correlation of shared loci across trait specific PRS may also reveal specific regions of the genome that exhibit pleiotropy for those traits, and potentially improve the localization of causal variants within these loci. Furthermore, genetics of behavioral traits could be explored with the use of multiple PRS, and possibly lead to detangling the environmental contribution of these behavioral traits on phenotypes of interest from the shared underlying genetics. Further methods development will be required to achieve this goal.

We have demonstrated that PheWAS, using a CRC PRS as a genetic score, can aid in detecting genetic correlation between CRC risk and other traits recorded as ICD codes in the medical record. This provides an orthogonal method to MR and LD-score regression. MR assumes all the SNPs have effect sizes in the same direction and that any direct pleiotropic effects of the SNPs on the outcome are distributed independently of the genetic associations with the risk factors. Neither of these assumptions are necessary for PheWAS with a PRS. LD-score regression typically uses summary statistics and requires harmonization of phenotypes and statistics across datasets. PheWAS with a PRS allows for the use of individual-level data that can be found in large biobanks as well as local clinics. Further analysis with other PRS is necessary to confirm the utility of PRS PheWAS as a reliable orthogonal method to detect genetic correlation between other traits.

## Web Resources

Phecodes: http://www.phecode.org

## Statements and Declarations

## Acknowledgments

This research has been conducted using the UK Biobank Resource under Application Number 47377. This work uses data provided by patients and collected by the NHS as part of their care and support.

eMERGE Network (Phase III): This phase of the eMERGE Network was initiated and funded by the NHGRI through the following grants: U01HG008657 (Group Health Cooperative/University of Washington); U01HG008685 (Brigham and Women’s Hospital); U01HG008672 (Vanderbilt University Medical Center); U01HG008666 (Cincinnati Children’s Hospital Medical Center); U01HG006379 (Mayo Clinic); U01HG008679 (Geisinger Clinic); U01HG008680 (Columbia University Health Sciences); U01HG008684 (Children’s Hospital of Philadelphia); U01HG008673 (Northwestern University); U01HG008701 (Vanderbilt University Medical Center serving as the Coordinating Center); U01HG008676 (Partners Healthcare/Broad Institute); U01HG008664 (Baylor College of Medicine); and U54MD007593 (Meharry Medical College).

## Competing Interests

The authors have no relevant financial or non-financial interests to disclose.

## Author Contributions

All authors contributed to writing and editing this manuscript. Data analysis and the first draft were done by Elisabeth A Rosenthal. All authors commented on previous versions of the manuscript. Study conception and design were by EAR, DRC, UP, and GPJ. Data curation was done by W-QW, YL, BN-K, ML, LK, JAP, DRC, and GPJ. Funding, resources and supervision were provided by DRC and GPJ. All authors read and approved the final manuscript.

## Data Availability

eMERGE stage 3 phenotype and genotype data are available in dbGAP under accession number phs001584.v2.p2. UKBB data is available at the UK Biobank Resource.

## Ethics approval

Ethics approval for the eMERGE stage 3 study was provided by each participating institution’s institutional review board.

## Consent to participate

Informed consent was obtained from all individual participants included in the study.

## Notes

### Competing Interest Statement

The authors have declared no competing interest.

### Author Declarations

IRB of Group Health Cooperative/University of Washington gave ethical approval for this work. IRB of Brigham and Women's Hospital gave ethical approval for this work. IRB of Vanderbilt University Medical Center gave ethical approval for this work. IRB of Cincinnati Children's Hospital Medical Center gave ethical approval for this work. IRB of Mayo Clinic gave ethical approval for this work. IRB of Geisinger Clinic gave ethical approval for this work. IRB of Columbia University Health Sciences gave ethical approval for this work. IRB of Children's Hospital of Philadelphia gave ethical approval for this work. IRB of Northwestern University gave ethical approval for this work. IRB of Boston Children's Hospital gave ethical approval for this work. IRB of Marshfield Clinic Research Foundation gave ethical approval for this work. IRB of Icahn School of Medicine at Mount Sinai gave ethical approval for this work.

