## Supplemental Information for "Mixed-effects polygenic risk score Phenome-wide association study detects genetic correlation between colorectal cancer risk and phenotype data extracted from the electronic health record"

Estimating pairwise kinship in emerge3 105K dataset

The Electronic Medical Record and Genomics (eMERGE) network has enrolled more than 105,000 participants through many sites, nationwide (Stanaway et al. 2019). The data set represents multiple ancestries, including admixed individuals, and some of the participants may be closely related. In order to analyze this dataset, the relatedness among close relatives, as well as the ancestral backgrounds of participants, must be accounted for to avoid bias and to reduce type 2 error. We calculated unbiased estimates of relatedness among participants in the eMERGE 105K dataset, using the model free method PC-relate (Conomos et al. 2016). In brief, the method estimates relatedness while adjusting for the presence of admixture, by partitioning the principal components of ancestry (PCs) into components due to distant ancestral population membership and components due to recent familial ancestry.

### Methods

For PC calculation, the number of SNPs was pruned by restricting pair-wise correlation (r^2^ ) < 0.2, reducing the number of SNPs by 88%, to 120,090. As there were over 105K participants, the number of participants was reduced to make the process computationally tractable. The method to reduce the number of participants ensured that all ancestral backgrounds were represented and that only unrelated individuals were removed from the calculation. These removed individuals were later projected onto the PC space (see step 6).

The following steps were used to estimate the values of kinship $(\theta$) and identities by state ($\varphi_{2}$,$\varphi_{1}$,$\varphi_{0}$) between pairs of individuals, where Kth degree or closer relatedness is defined as $\theta> {\frac{1}{2}}^{(K+1+\frac{1}{2})}$. The addition of 1/2 in the exponent is used to account for possible error in the estimate of kinship ($\hat{\theta}$), allowing for an increase in the number of potentially related pairs.

1. Calculate initial estimates of relatedness ($\hat{\theta_{0}}$) among all pairs of participants using KING method of moments (Manichaikul et al. 2010).
2. Separate the participants into a set containing related individuals (RSet) and a set containing unrelated individuals (USet) using the function pcairPartition() from the GENESIS package (Gogarten et al. 2019), where relatedness is defined by 4^th^ degree or closer (K=4). The resulting sets contained N=10,608 (Rset) and N= 94,500 (Uset) participants. Note the selection of participants in the USet by the GENESIS pcairPartition() function optimizes the representation of all ancestries.
3. To reduce computation time and memory requirements, a random 20% sample from the USet was taken (N= 18,900). However, some of the participants in this list may be related to those in the RSet. Of the 80% of USet that were not selected, N=4,884 were related to at least one participant in the RSet at K=4 degrees or closer. These 4,884 participants were added back to the random 20% resulting in 23,784 unrelated participants in the pruned USet list.
4. Estimate initial PCs of ancestry using participants in the pruned USet using the function snpgdsPCA() from the package SNPRelate (Zheng et al. 2012). Project these PCs onto the related participants. The first four PCs explained 4.2% of the variation in the pruned USet and are used in step 5.
5. Estimate the updated kinship estimates, $\hat{\theta}_{1}$, among the RSet and pruned USet, using the function pcrelate() from the GENESIS package and the first four PCs from step 4.
6. Estimate revised PCs using participants from both the RSet and pruned USet using the pcair() function from the GENESIS package, adjusting for $\hat{\theta}_{1}$ calculated in step 5. Project these PCs onto the participants excluded from the pruned USet.
7. Calculate the final estimate of kinship ($\hat{\theta}$) and identities by state (IBS) ($\hat{\varphi_{2}}$, $\hat{\varphi_{1}}$, $\hat{\varphi_{0}}$) with the pcrelate() function from the GENESIS package, using the first four revised PCs from step 6.

Relatives, defined by K=3 degrees of relatedness were then classified into three groups: duplicates, dyads, and clusters. Duplicates consist of participants that are either monozygotic (MZ) twins or duplicates of the same individual’s sample. These are defined by { $\hat{\theta}> {\frac{1}{2}}^{(1+\frac{1}{2})}$ AND $\hat{\varphi_{0}}$ < 0.2 AND both participants have the same sex and birth year}. Only a single participant from each of these duplicate pairs was retained in further analysis. Dyads are defined as pairs where each participant is present in only one pair, after removing the duplicate pair set. Clusters are defined as three or more participants that are related within 3 degrees to at least one other person in that cluster.

### Results

Of the initial 105,108 participants, there were 13,546 participants found to be in at least one of 11,260 related pairs. Ninety-two pairs were determined to be duplicates or MZ twins. There were 4,477 dyads of 2 related samples, including four in which one of the participants was part of a duplicate pair. There were 1,012 clusters of more than 2 related samples, containing a total of 4,426 participants, including seven that contained a duplicate participant.

Code

Code for PC and kinship calculation is available upon request at https://github.com/earosenthal/relatedness-calculations

Checking for bias in effect estimates

We performed a mixed model PheWAS in the UKBB cohort, excluding CRC cases for all phecodes that replicated in the eMERGE cohort. If the effect size changes substantially, then the presence of the CRC cases in the original analysis may be biasing the estimated effect. This bias may be due to several factors. It is possible that the phecode is likely a downstream result of CRC or its treatment, which we failed to remove from the analysis. It is also possible that some of the observed association is due to non-genetic factors. This is particularly relevant for modifiable risk factors that are known to be correlated with CRC, such as smoking, obesity, and alcohol consumption.

### Methods

We removed all participants who were considered cases for CRC (phecode 153) in the United Kingdom Biobank (UKBB) discovery cohort and performed the mixed model PheWAS as described in the main paper. We calculated the percent change in effect size estimate from using the entire cohort.

### Results

Results are given in the Supplemental Table 2. The effect estimates for Septicemia and Acquired absence of breast reduced by approximately half. For the remaining eight phecodes, there was little change (< 20%). Interestingly, the effect size estimate for Cancer of prostate increased slightly (4.7%). The effect sizes for Alcohol-related disorders and Tobacco use disorder were virtually unchanged.

### Tables

Supplemental table 1: Results from the mixed-model PheWAS analysis in the discovery cohort (UKBB) and replication cohort (eMERGE). Significance is determined by the Score test. Columns are stored in general mode so that small p-values are not replaced with zero. In the phecode column, the leading zero is replaced with a capital letter O. A ROW column is included to help with sorting. Other columns are defined as follows: ancestor=1 if the phecode is an ancestor code; consequence=1 if the phecode is a consequence of CRC diagnosis or its treatment as determined by William M. Grady; Score=value of the score function; Score.SE=estimated standard error of Score; Score.stat=Score Z test statistic; p=p-value; beta = estimated effect size for the phecode; SE=approximate standard error of beta; PVE=approximate proportion of PRS variance explained by the phecode.

**Supplemental Table 2: Comparing the effect estimates (β), rounded to two decimals, from the discovery cohort (UKBB) with and without CRC cases.**

| **Phecode** | **Description** | **Discovery β** | **CRC cases Removed β** | **% change** |
| --- | --- | --- | --- | --- |
| 208 | Benign neoplasm of colon | 0.25 | 0.22 | -9.4 |
| 175 | Acquired absence of breast | 0.05 | 0.02 | -53 |
| 1100 | Family history | 0.04 | 0.03 | -9.5 |
| 317 | Alcohol-related disorders | 0.04 | 0.04 | -0.13 |
| 174 | Breast cancer | 0.03 | 0.02 | -12 |
| 211 | Benign neoplasm of other parts of digestive system | 0.05 | 0.04 | -19 |
| 185 | Cancer of prostate | 0.02 | 0.02 | 4.7 |
| 038 | Septicemia | 0.03 | 0.01 | -45 |
| 278 | Overweight, obesity and other hyperalimentation | 0.02 | 0.01 | -15 |
| 318 | Tobacco use disorder | 0.02 | 0.02 | -1.3 |
